# The Experience of Two Independent Schools with In-Person Learning During the COVID-19 Pandemic

**DOI:** 10.1101/2021.01.26.21250065

**Authors:** Darria Long Gillespie, Lauren Ancel Meyers, Michael Lachmann, Stephen C Redd, Jonathan M Zenilman

## Abstract

**BACKGROUND:** In 2020, U.S schools closed due to SARS-CoV-2 but their role in transmission was unknown. In fall 2020, national guidance for reopening omitted testing or screening recommendations. We report the experience of 2 large independent K-12 schools (School-A and School-B) that implemented an array of SARS-CoV-2 mitigation strategies that included periodic universal testing.

**METHODS:** SARS-CoV-2 was identified through periodic universal PCR testing, self-reporting of tests conducted outside school, and contact tracing. Schools implemented behavioral and structural mitigation measures, including mandatory masks, classroom disinfecting, and social distancing.

**RESULTS:** Over the fall semester, School-A identified 112 cases in 2320 students and staff; School-B identified 25 cases (2.0%) in 1200 students and staff. Most cases were asymptomatic and none required hospitalization. Of 69 traceable introductions, 63(91%) were not associated with school-based transmission, 59 cases (54%) occurred in the 2 weeks post-Thanksgiving. In 6/7 clusters, clear noncompliance with mitigation protocols was found. The largest outbreak had 28 identified cases and was traced to an off-campus party. There was no transmission from students to staff.

**CONCLUSIONS:** Although school-age children can contract and transmit SARS-CoV-2, rates of COVID-19 infection related to in-person education were significantly lower than those in the surrounding community. However, social activities among students outside of school undermined those measures and should be discouraged, perhaps with behavioral contracts, to ensure the safety of school communities. In addition, introduction risks were highest following extended school breaks. These risks may be mitigated with voluntary quarantines and surveillance testing prior to re-opening.

## BACKGROUND

SARS-CoV2 is a novel, highly infectious virus spread via respiratory droplets. It first appeared in December 2019 and has since spread to every country in the world. Infections have been particularly virulent in the United States, with 19.2 million cases and >398,000 deaths as of January 20, 2021 ^1^.

The growing pandemic led to school closings throughout the country beginning in March 2020. Large numbers of children were rapidly transitioned to virtual instruction marking the greatest challenge to the US educational system since the 1918 Spanish flu pandemic. One justification for the closings was concern that children are more likely to contract and spread respiratory viruses in a congregate setting. However, research showed that schools played a minor role in previous SARS and MERS-CoV outbreaks.

Although we learned much about the virus in the ensuing months, it remained unclear how opening schools would affect transmission. Nonetheless, given the continued high levels of infection in the US, most school districts either delayed in-person education for the first semester of the 2020-21 school year or opted for a hybrid approach.

During summer 2020,, many political and health officials called for schools to reopen^3^ In March 2020, the Centers for Disease Control and Prevention (CDC) posted guidelines for managing school operations during the pandemic, updating them in August 2020 with a section on the benefits of in-person schooling. Notably missing was any guidance on testing^4^ and CDC did not release testing recommendations for schools until December 2020^5^ While state and local health authorities provided technical guidance consistent with national recommendations, they were ill-equipped to provide specific advice on school-based testing. Furthermore, absent a comprehensive national testing strategy and shortages of test kits,, they could not provide resources to implement such testing. These circumstances left local school officials on their own to determine reopening plans, including laboratory testing, and to identify the financial and laboratory testing resources to implement such testing.

We report here on the experience of two independent K-12 schools, one in the Southeast and one in the Mid-Atlantic, and their collaborative efforts to reopen for the 2020 fall semester. Each school followed the CDC’s guidelines designed to prevent the transmission of SARS-CoV-2 while also including aggressive laboratory screening similar to that which colleges and universities were implementing^6^. A critical aspect to the reopening was full transparency and open communication with parents, staff, and students. The schools also recognized that reopening would require continual reassessment and adjusting to changing community epidemiology and resources.

We report our experience to identify the challenges and principles that policy makers and educational leadership could follow to return to in-person learning while protecting students and staff.

## METHODS

### Participants

Administrators at 2 independent, K-12 schools located in the South (School A) and Mid-Atlantic (School B) allowed the investigators to monitor their experiences as they returned to in-person learning during the fall semester of 2020. School A had 2299 students and staff; School B had 1200. Their reopening plans differed slightly, but both were based on 4 major elements (Table 1): stakeholder engagement, physical infrastructure, policies and operations, and laboratory screening and testing of students and staff for SARS-CoV-2. Both plans were developed in consultation with medical advisors.

**Table 1.** Interventions undertaken to sustain in-person education, School A and School B.

|  | School A | School B |
| --- | --- | --- |
| Stakeholder Engagement + Activities |  |  |
| Community contract | • Avoid large gatherings<br>• Quarantine the entire family if 1 member tests positive for SARS-CoV-2<br>• Report COVID-19 symptoms proactively to the school<br>• Reduce in-community activities<br>*Implied contract; no signature required | Agreement electronically signed by parents |
| Physical Infrastructure |  |  |
| Desks | Changed from group tables to desks 6 feet apart. | Most spaced 6 feet apart, some 5 feet apart. |
| Schedule | Staggered class starts to reduce queuing | Shifted to block scheduling for middle and upper school students (4 classes per day) with 15 minutes of passing time between each class |
| Restrooms | Limited number of people in restrooms based on number of stalls/urinals | Limited number of people in restrooms based on number of stalls/urinals. |
| Hand sanitizer stations | • Sanitizer placed at entrance to every building and class.<br>• Outdoor hand-washing stations. | Sanitizer placed at all major entrances, division offices, and common areas. |
| Disinfection | Teachers disinfected each desk and doorknob between classes. | Classrooms disinfected between each class for middle- and upper-school students. Classrooms disinfected with R-Zero UV light system at the end of the day. |
| Airflow | • MERV filters enhancement | • MERV-13 filters enhancement |
|  | • Dedicated outdoor air systems opened to maximum<br>• Bipolar deionization in mechanical system | • Windows and doors in classrooms open<br>• Windows on buses open |
| Temperature Checking | Kogniz IR cameras at main entrances | Kogniz IR cameras at main entrances |
| Changes in policies and operations |  |  |
| Carpool drop off and pick up | Social distancing at dropoff | Social distancing at dropoff |
| Mask wearing | Double-layer mask required at all times except for snack and lunch, which are consumed silently at desks, and for a daily outdoors, socially distanced, 5-minute break. | Required at all times, removed only for eating/drinking. |
| Participation in extracurricular sports | Many extra-curricular activities cancelled (particularly for lower school)<br>• Varsity sports continued, but with regular PCR and rapid testing of all players; small pod training; | Extra curricular activities cancelled. |
| Conversation during lunch | • No conversation while eating.<br>• Movies shown during lunch to limit talking. | Limited in classrooms; allowed if eating outside. |
| Congregating activities | No congregate activities | Middle- and upper-school chapel held via Zoom. No gatherings greater than 25 allowed. |
| Quarantine policies | • Positive child isolated for 10 days<br>• siblings of positive child isolated for 14 days<br>• | • 14 days off campus after positive test.<br>• Family of student must remain off campus for additional 14 days. |
| Contact tracing | - Positive child isolated for 10 | Contact tracing via Excel |
| procedures | days from test<br>- siblings of positive isolated<br>for 24 days from sibling test | spreadsheet |
| Remote learning triggers |  | Based on consultation with medical advisors. |
| In-school sports | All sports continued with protective protocols and enhanced testing over holidays and before events. | Conditioning only for most of semester; intramural basketball games with players masked began in early December. |
| Out-of-school sports | No school position | Discouraged in community contract. |
| Testing procedures |  |  |
| Test type | ● Round 1: PCR, anterior nares collection<br>● Round 2: Nasopharynx PCR for upper school, Thermo Fisher TaqPath saliva test for lower- and middle-school students<br>● Round 3 and ongoing: pooled saliva PCR via SalivaDirect™ | ● Rounds 1 - 5: Thermofisher TaqPath assay, anterior nares collection<br>● Rounds 6-16: SalivaClear saliva collection |
| Frequency of universal laboratory testing | At entry; monthly while doing individual tests; weekly once switched to pooled saliva testing | Half the population every week. |
| Individual screening in addition to universal screening | ● Contacts of positive cases<br>● Athletes<br>● Symptomatic individuals | ● Contacts of positive cases<br>● Athletes |
| Sewage testing | None | Twice weekly |

Both schools had extensive contingency plans to close in-person learning and transition to virtual education if needed. Key metrics used to determine whether to invoke these contingency plans included community positivity rate, in-school positivity rate, and presence of in-school transmission. In-school transmission was a key differentiator, because school leadership acknowledged that introductions related to community exposures outside school would occur at community rates - but a key element was making certain that schools did not play a role in amplifying cases, i.e. limiting in-school transmission, and closing classrooms or perhaps entire sections of the school, or changing protocols if in-school transmission started to climb.

### SARS-CoV2 Testing

Laboratory screening protocols differed slightly by school and evolved over the course of the fall semester based on testing options, availability, and logistics. Schools initially collected nasopharynx samples from the entire student body and staff at biweekly or monthly intervals depending on schedules and testing capacity. Each universal testing event required 2-3 days to implement and to obtain all results. As the semester progressed, both schools transitioned from nasal swab PCR to saliva-based tests by a vendor using the SalivaDirect™ Yale protocol^7^, enabling more frequent testing..

Pooling of saliva specimens was performed with up to 24 specimens per pool. If a pool was positive, individual samples were reflex tested either by SalivaDirect™ or a confirmatory PCR, with results classified as positive, negative, or inconclusive within 48 hours of collection. School officials then notified test participants of the results. A case was defined as a person with a positive laboratory test for SARS-CoV-2.

For positive cases, school officials implemented contact tracing in conjunction with local health authorities to determine the likely source of exposure. The infected person and those within close proximity or where a potential breach of protective measures could be determined were tested, quarantined, and asked to quarantine and report any symptoms.

As community rates rose in the fall, the schools tested specific populations such as athletes more frequently. They also tested the entire school population after the Thanksgiving break. Officials also encouraged parents to report any test results obtained outside the school.

### Case Definitions and Exposure Estimation

Positive cases were identified as self-reported (SR) if the person was symptomatic or had a known contact and was tested at school or independently; universal test (UT), if the person was asymptomatic and tested as part of regular screening; or converted during quarantine (CDQ), if the person was identified as a known contact due to UT or contact tracing and was quarantining when they received a positive test. We used a cluster analysis for each case to identify common linkages, source of introduction, and potential route of transmission.

To calculate the rate of asymptomatic versus symptomatic cases, we identified all cases with symptoms detected through SR, UT, or CDQ and followed all asymptomatic individuals identified, and asked individuals or their families to report any symptoms. We then calculated the proportion of symptomatic cases by age cohort. Severe cases were defined as infected persons requiring hospitalization. We defined a cluster as any event which involved 2 or more linked cases in school.

Using data from UT and SR, we calculated the rate of virus introduction into each school and the rate of transmission per introduction. To accurately determine the number of introductions into the school setting, we excluded students from testing before school started (Round 1), as those students did not enter the school and were hence not introductions. Self-reported cases occurring more than 1 week before UT; all cases affiliated with an off-campus, non-school sanctioned event over Thanksgiving break; all secondary cases; and all self-reported cases occurring after the last round of UT. This likely significantly underestimates transmission, since we only analyzed cases with tracking data.

### Data Analysis

We grouped those who tested positive into clusters based on the site of exposure (family, community, or school). We then calculated the average reproductive rate as the number of secondary cases generated by contact with the infectious person.

We calculated the binomial confidence intervals (CI) using Jeffrey’s interval. We used a CI with accepted multipliers for incidence based on local seroprevalence to compare in-school versus community cases.

We calculated the average number of infections related to the initial infectious individual to determine the reproduction number of the virus in the school community^8^, *R*_*0*_. Here we assume that each outbreak represents only one generation of transmission. Since the overall rate of transmission, conditional on infection, is low, this assumption is valid (assuming that secondary infections follow the same low rate). With this assumption the estimated rate of spread will slightly overestimate the real rate. We used the maximum likelihood estimator for negative binomial distribution of the outbreak sizes to estimate the overdispersion parameter *k*. Our model also used the viral reproduction rates to assess the relationship between testing frequency and delayed detection on the potential number of cases. All analysis was done using the R statistical computing environment^9^.

## RESULTS

School A reported 109 confirmed COVID-19 cases (4.9% of students, faculty, and staff) between August 5, 2020 and December 20, 2020 (Table 3). Sixty cases (54%) were identified through the 9 rounds of UT; 22 (20%) through contact investigation based on UT-identified cases; and 30 (27.8%) self-reported. Eleven (10%) of cases in school A were identified during the initial testing (n=11), and 59 (54%) were identified during the 3 weeks following the Thanksgiving break.

School B reported 25 confirmed COVID-19 cases (2.0% of students, faculty, and staff) between August 24 and December 20. Twenty-one cases (84%) were identified through UT; 1 (4%) through contact investigation resulting from a UT case; and 3 (12%) were self-reported. In both schools, cases peaked in the period after school breaks (summer holiday, fall break, and Thanksgiving break) (Figure 2). The detailed tables of testing results in both schools are available in the Supplemental Material. Adults in both schools were more likely than lower school students to demonstrate symptoms at the time of testing (47.6% vs 2.6%) (Table 2). No cases require hospitalization.

**Table 2.** Proportion of Laboratory-Confirmed Cases That Were Symptomatic At Time of Testing Combined Schools A and B

|  | Total Infections | Total Symptomatic | Symptomatic Rate | 95% Confidence Interval |
| --- | --- | --- | --- | --- |
| Lower School | 39 | 1 | 2.6% | 0.3-11.1% |
| Middle School | 20 | 5 | 25% | 10.2-46.4% |
| Upper School | 57 | 5 | 8.8% | 3.4-18.2% |
| Adults | 21 | 10 | 47.6% | 27.7-68.1% |

**Table 3.**
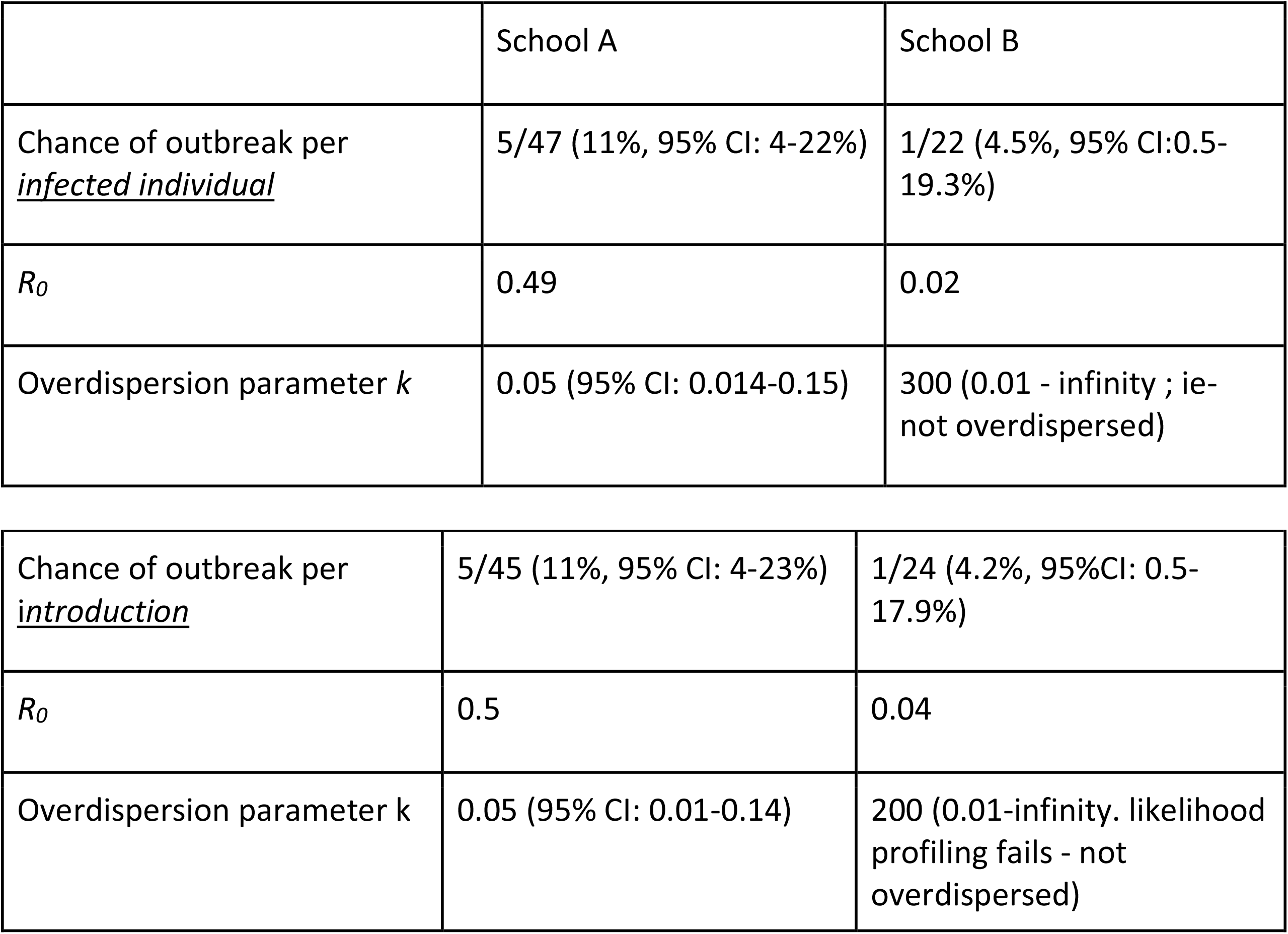
Characteristics of outbreaks in School A and School B.

**FIGURE 1.**
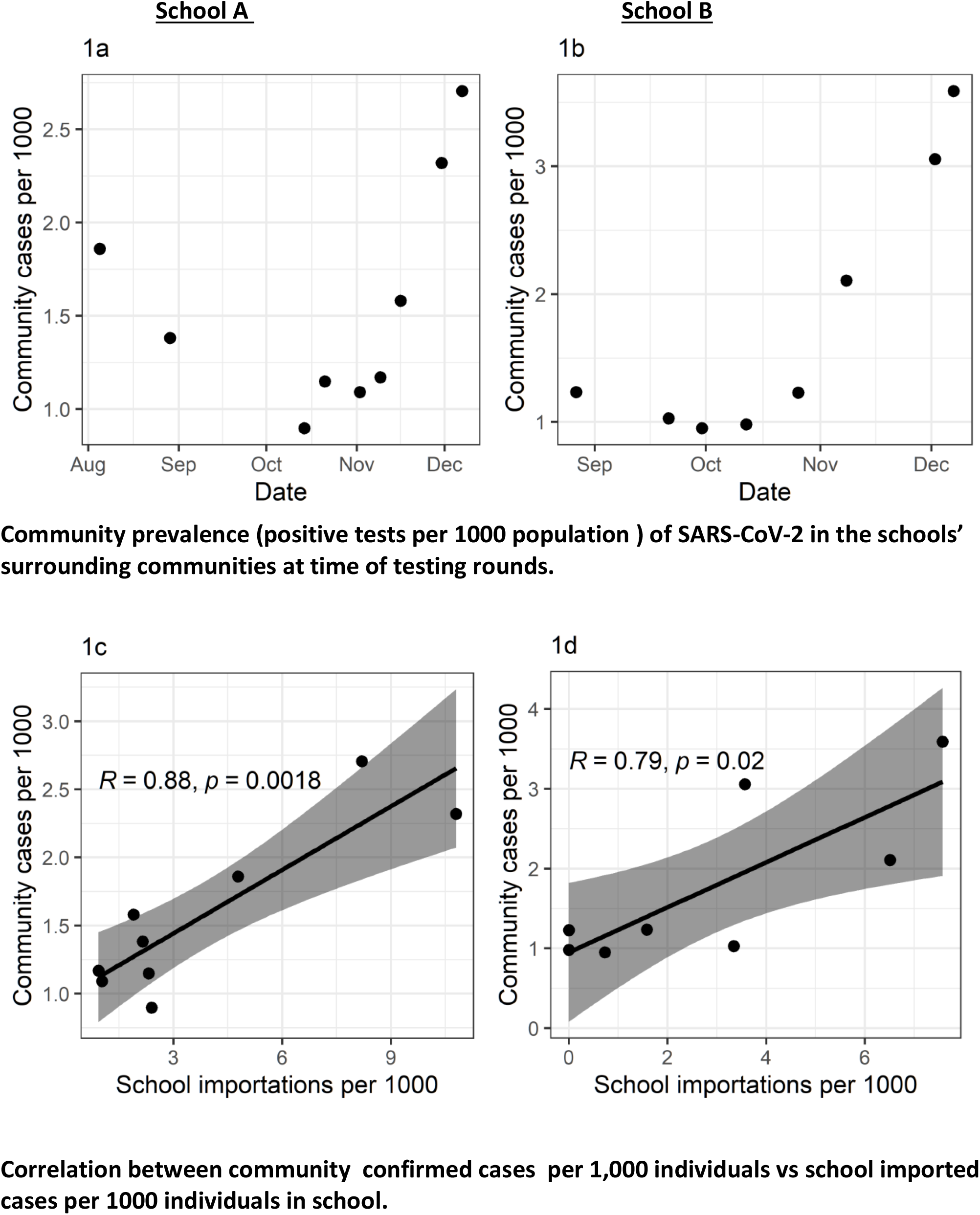
Community Incidence compared with school introductions.

**Figure 2A.**
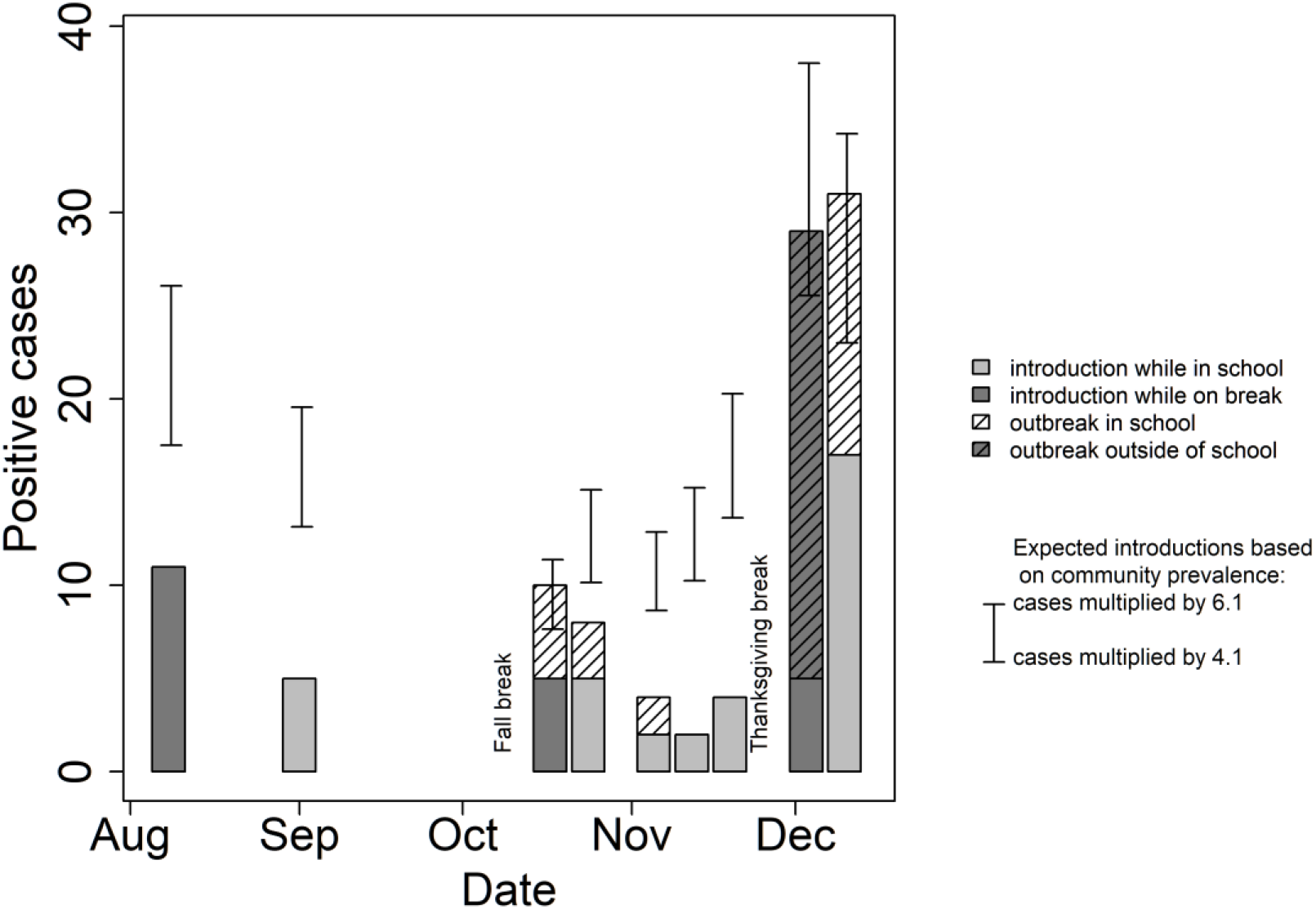
Case Incidence –School A.

**Figure 2B.**
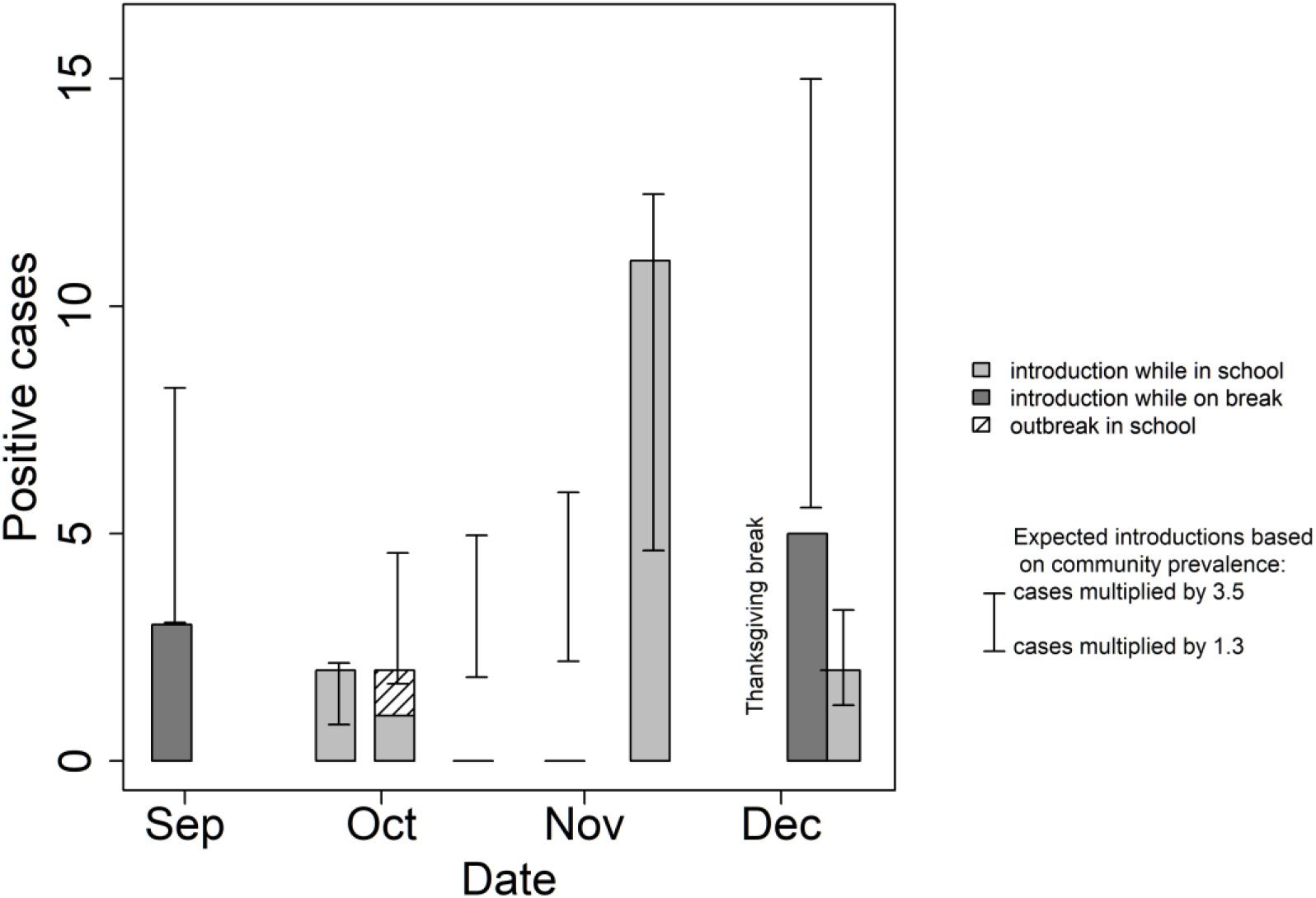
Case Incidence—School B.

Differences between the schools that may account for disparities in testing results include size of the student and staff testing; testing frequency; and continuance of sports, since the largest outbreak at School A was linked to a non-school sanctioned, sports-related event.

### Community incidence rates

As Figure 1 shows, community incidence rates correlated with school infections (Pearson correlation 0.9, p<0.01 for School A; 0.8, p< 0.05 for School B) There was no correlation between community positivity rates and in-school introduction (School A, correlation 0.57, P>0.1; School B: correlation 0.27, P>0.1). Using multipliers based on contemporaneous seroprevalence studies, we found that in-school rates were consistently below community infection rates.

### In-school transmission

Nine percent of the 69 introduced cases in both schools transmitted the virus (Figure 3). Five secondary infections from the 45 introduced cases occurred in School A; 1 secondary infection from the 24 introduced cases occurred in School B. The risk of an outbreak per introduction and per infected individual are described in Table 3, and the R_0_ (reproduction number) was consistently low; 0.47 in School A and 0.05 in School B.

The outbreak clusters are described in more detail in Table 4. Of the 6 outbreaks observed in school A, the minimum number of secondary cases was 3. The relative lack of transmission singletons, but many non-transmission events suggests an overdispersed chance of transmission in School A. School B was not overdispersed, since the only outbreak had a single transmission event - the smallest dispersion possible, although we did not have the statistical power to conclude this definitively. There was no evidence of student-to-teacher or teacher to student transmission in either school.

**Table 4.** Epidemiologic Characteristics of Clusters Identified

Epidemiologic Characteristics of Clusters Identified
|  | School A |  | School B |  |
| --- | --- | --- | --- | --- |
|  | Positive cases | Potential infectious event | Positive cases | Potential infectious event |
| Cluster 1<br>in-school transmission among lower-school students | 6* | Close proximity in classroom | 3 | A younger sibling student had been symptomatic and stayed home; the older sibling was tested in universal testing and found to be positive. A younger sibling then tested and was found to be positive. A classmate of the younger sibling was found to be positive on the next round of weekly testing |
| Cluster 2<br>in-school transmission among upper-school students | 7** | Close proximity in cafeteria |  |  |
| Cluster 3<br>off-campus transmission, 15 individuals were confirmed as cases in universal screening; contact tracing identified that they had all attended the same off-campus independent party. An additional 13 cases were confirmed as cases, 3 during school directed quarantine and 6 through self-reported, and 4 more discovered in the following weekly round. (28 cases, 25% of total) | 28 | Off-campus party |  |  |
| Cluster 4<br>universal laboratory screening, 4 individuals in one lower school class were confirmed as cases and an additional person had an inconclusive test result. On repeat testing, the student with the inconclusive test result was confirmed as a case. Repeat testing then found two additional students in the same class infected and one sibling who converted in quarantine. | 8 | In-classroom transmission |  |  |
| Cluster 5<br>3 individuals confirmed as cases on universal testing in one lower school class, with an additional student who was initially negative on universal testing, on repeat testing found to be positive. The class was quarantine, and two additional students converted to positive in quarantine | 6 | Unknown |  |  |
| Cluster 6<br>1 student was identified on self-report testing after a possible out-of-school exposure, and reported to the school. Two other students were later found to be positive on self-report. These students had contact with each other off-campus, as well | 3 | Unknown, likely off-campus socializing |  |  |

### Infection source

Seventy two percent of in-school transmission cases in School A were associated with noncompliance with school mask wearing rules. Of known off-campus sources, the major ones identified were family exposure, including siblings returning from college; off-campus activities, including parties and other gatherings. However, the source of the majority of infections could not be determined.

## DISCUSSION

This study evaluated the experience of two large independent K-12 schools which implemented in-person instruction accompanied with multifaceted SARS-CoV2 mitigation strategies which included universal periodic testing of all students and staff. Both schools were able to successfully maintain in-person schooling for the full fall 2020 semester, despite rising community numbers and other neighboring school closures.

There were 4 key findings from this study. First, while school-conducted universal testing showed that children can contract and spread SARS-CoV-2, the majority of cases these schools identified did not lead to larger chains of transmission.. Indeed,, with the mitigation measures implemented there was a significantly lower transmission rate than seen, without such measures, as has been seen in previous respiratory virus seasons with other viruses such as influenza^10^.

These data are consistent with recent reports from Europe which also found that school-based transmission is low^11,12^. The occurrence of many outbreaks when protocols were not fully followed shows how important these measures are for keeping transmission rates low.

Second, we found no correlation between in-school infections and community positivity rates, which calls into question the use of community positivity rates as a metric for school openings and closings. However, we did find a correlation with actual community incidence.

Part of this may represent the realities of community testing. In our schools, the school, positivity rates (positives/total number of tests) and case incidence (positive tests /population) are the same, because everybody is tested. In the community they are different, because only a small proportion of the community are being tested In-school introductions did correlate well with community cases per 1,000 population.

Third, we also found that the distribution of secondary infections (transmission) is bimodal, with 91% of identified cases having no secondary transmission, and 9% of introduced cases accounting for all the identified clusters. The majority of clusters occurred when mitigation protocols were not followed, whether in or out of school. When secondary transmission occurred, the resulting clusters in School A included at least 5 additional cases. These clusters might be considered small “super spreader events” and are consistent with other studies of SARS-CoV-2 transmission. They also highlight the effectiveness of mitigation protocols in preventing transmission.

Finally, based on the chronology of infections, the rate of positive cases was highest following school breaks and when there was clear evidence of students attending social or family events in the community, and lowest when in-person schooling continued uninterrupted. Indeed, in-school infections peaked each time students returned from a prolonged school break.

The schools’ testing protocols had additional benefits. They served as a gauge of the effectiveness of mitigation protocols when adhered to; identified risky activities; enabled school officials to adjust protocols and processes in real time; and provided reassurance for families and faculty. Although we believe that our identification of individual sporadic cases prevented the further development of case clusters, this cannot be quantified with the data we have available.

We found that temperature screening was not useful, because nearly all cases identified were asymptomatic. School B also implemented SARS-CoV2 sewage surveillance, however, results from that activity were not useful in real-time in identifying new case incidence due to delays in obtaining results as well as the difficulty in interpreting positive results in a 1200 person school community.

Methods to further identify the role of schools in viral transmission include surveillance screening in schools during periods of more intense community transmission and outbreak investigations in schools with assessments of secondary household transmission. Until this work is complete, it would be premature to draw definitive conclusions on the importance of schools as part of community transmission.

### Limitations

Our report has 2 major limitations. First, our data reflect the experience of 2 schools that were able to invest substantial financial, logistical and organizational resources in a testing program. We recognize that these resources are not available to the vast majority of institutions. However, our goal was to assess the experience of these schools and the importance of regular testing in safely returning children to school.

The second major limitation is the adaptive testing protocol used over the 4-month period, in which the protocol changed in real time based on availability of testing resources. The schools initially began with less frequent testing than recommended in settings with a high risk of transmission^13^. Modeling results showed that school transmission might have gone undetected with the lower frequency of testing. However, when less expensive and easier-to-implement pooled saliva testing became available, each school increased its testing frequency. This design, wherein each round of testing was used to decide how frequently to repeat the screening process, was sufficient to confirm the risk of introductions and school transmission.

Our modeling estimates are limited, since the number of outbreaks was small and we were only able to identify one generation of infection. Therefore, our estimate of *R*_*0*_ is an upper bound. Nevertheless, we found that our R_0_ estimates were consistently substantially below 1, indicating that in-school transmissions did not represent sustainable outbreaks.

## CONCLUSIONS

These results highlight that while SARS-CoV-2 is infectious in children, in schools which implemented a comprehensive strategy, transmission can be controlled. Testing is a key tool in identifying asymptomatic cases., with testing being a key tool in the arsenal. Our data highlights the challenge of asymptomatic infections, out-of-school social activities, violation of face mask rules, and return to school after extended breaks.

Given that the vast majority of our youngest students were asymptomatic at time of testing, this presents an additional challenge to schools and requires strict adherence to protective protocols. However, it does bear highlighting that few cases occurred in staff, and that these all seemed to have had out-of-school introduction sources. Also, there were no severe cases among students or staff. Given that both schools are well-resourced, with a population that likely has a lower burden of chronic disease and better access to medical care, the exact consequence of these introductions in less well-resourced communities is not known.

In addition, the greatest vulnerability for transmission was a lack of protocol adherence in school and during off-campus parties, particularly around the holidays. As schools open, awareness of these key vulnerability points will be crucial.

These results suggest that the transmission of the SARS-CoV-19 virus between children and adults may differ from influenza virus transmission. In primary and secondary schools with comprehensive infection mitigation programs SARS-CoV-19 transmission can be either be prevented or managed effectively, and may represent a different epidemiology compared to colleges and universities^14^. However, we recognize that because these institutions are residential settings, they differ substantially from the typical K-12 school.

The experience of these 2 independent K-12 schools show that in-person schooling can be conducted relatively safely, but not risk free, even in areas with moderate community COVID-19 incidence, if structural and behavioral mitigation strategies, augmented by aggressive testing, are implemented.

These results should encourage educational decision makers to assess their ability to provide periodic laboratory screening as way to identify virus infections and limit in-school and community transmission.

However, to bring this level of testing and behavioral change to schools with fewer resources will require that educators and public health officials provide significant financial and logistical support to enable schools to institute these protocols.

## IMPLICATIONS FOR SCHOOL HEALTH

The experiences of these 2 schools show that structural and behavioral mitigation strategies together with aggressive testing can safely allow a return to in-person education in K-12 grades, even in areas with moderate community COVID-19 prevalence. Such an approach could also detect the beginnings of larger outbreaks that may require a switch to remote learning.

These findings are particularly important in the context of the growing consensus that in-person schooling is critical for educational and social development of children^3,15^.

Our findings also suggest that implementing universal screening and following mitigation measures could have a behavior-modifying and protective benefit for students, school staff, and the community. Whether testing can serve an adjunctive role in ensuring adherence to mitigation measures seems likely but cannot be definitively determined.

Thus, there is a critical need for educational and public health support of rapid expansion of school-based testing capacity and the resources required if communities are to return to inperson education. Public policies that support in-person education during the SARS-CoV2 pandemic need to recognize these issues, and support lesser resourced schools with the testing, financial, logistical, staffing, and training resources to safely return children to school.

### Human Subjects Approval Statement

This project was reviewed by the Johns Hopkins IRB and it was as Not Human Subjects Research, as it was program evaluation of the testing program undertaken as part of the schools’ ongoing efforts to sustain in-person learning. The analysis was supported by the leadership at both institutions. We did not collect or have access to names or other identifiable information.

## Data Availability

We will provide the data to any interested party

## ACKNOWLEDGEMENTS

We are indebted to the students, staff, and parents of the two schools.

