## Supplemental Material for "The Experience of Two Independent Schools with In-Person Learning During the COVID-19 Pandemic"

**SCHOOL A Laboratory test results, August 5 – December 20**

| | ROUND 1 | Self | ROUND 2 | Self | Convert | Round 3 | Self | Convert | Round 4 | Self | Convert | Round 5 | Self | Convert | Round 6 | Self |
|---|---|---|---|---|---|---|---|---|---|---|---|---|---|---|---|---|
| | (Before start) August 5 -10 | Aug 11-Sept 6 | Aug21-Sept11 | Sept 7-Oct 18 | | Oct 7 - 20 | Oct 19 - 25 | | Oct 21-28 | Oct 26 - Nov 1 | | Nov 2-4 | Nov 2 - Nov 8 | | Nov 9-12 | Nov 9 - Nov 15 |
| LS Students | 0 | 0 | 1 | 0 | 0 | 5 | | | 0 | 0 | 0 | 0 | 0 | | 0 | |
| MS Students | 2 | 1 | 1 | 2 | 0 | 0 | 1 | | 0 | 0 | 0 | 2 | 0 | | 0 | |
| US Students | 7 | 1 | 1 | 2 | 0 | 1 | 1 | | 3 | 1 | 1 | 0 | 0 | 1 | 2 | |
| Adults | 2 | 1 | 0 | 2 | 1 | 2 | 2 | | 0 | 0 | 0 | 0 | 0 | | 0 | 1 |
| Total Positives | 11 | 3 | 3 | 6 | 1 | 8 | 4 | 0 | 3 | 1 | 1 | 2 | 0 | 1 | 2 | 1 |
| Total Tests | 2299 | | 2320 | | | 2081 | | | 2158 | | | 1934 | | | 2136 | |

| Round 7 | Self | Convert | NO TESTING SCHOOL HOLIDAY | Self | Round 8 | Self | Convert | Round 9 | Self | Convert | Self | Convert | Total |
|---|---|---|---|---|---|---|---|---|---|---|---|---|---|
| | Week 14 | | | Week 15 | | Week 16 | | | | | | | |
| Nov 16-19 | Nov 16 - Nov 22 | | Nov 23 - 26 | Nov 23 - Nov 29 | Nov 30-12/3 | Nov 30 - Dec 6 | | Dec 7-11 | Dec 7-13 | | Dec 14-20 | | |
| 0 | | | | | 2 | 0 | | 8 | 0 | 0 | 3 | 6 | 25 |
| 0 | 1 | | | 0 | 0 | 1 | 1 | 0 | | 1 | 3 | 0 | 13 |
| 1 | 0 | | | 1 | 15 | 0 | 9 | 4 | 1 | 1 | 1 | 0 | 54 |
| 1 | 0 | 1 | 0 | 1 | 0 | 0 | | 0 | 2 | | 1 | | 17 |
| 2 | 1 | 1 | 0 | 2 | 17 | 1 | 10 | 12 | 3 | 2 | 8 | 6 | 112 |
| 2102 | | | | | 2687 | | | 2074 | | | | | |

**SCHOOL B Laboratory test results, School B August 24 – December 7**

| | Round 1 | Round 2 | Round 3 | Convert | Round 4 | Round 5 | Round 6 | Round 7 | Self | Self | Round 8 | Total |
|---|---|---|---|---|---|---|---|---|---|---|---|---|
| Week of: | 8/24/20 | 9/7/20 | 9/21/20 | | 10/5/20 | 10/19/20 | 11/2/20 | 11/16/20 | | | 11/30/20 | |
| Positives | | | | | | | | | | | | |
| LS (1-6) | 1 | | 2 | 1 | 1 | | | 4 | 1 | | 4 | 14 |
| MS (7-9) | | | | | | | | 1 | | | 3 | 4 |
| US (10-12) | | 1 | | | | | 2 | | | | | 3 |
| Adults | | 1 | | | | | 1 | | 1 | 1 | | 4 |
| Total Positives | 1 | 2 | 2 | 1 | 1 | 0 | 3 | 5 | 2 | 1 | 7 | 25 |
| Total Tests | 743 | 1156 | 1366 | | 1370 | 1448 | 1417 | 879 | | | 1666 | |

Laboratory test results, School B August 24 – December 7 (School B)

| | Round 1 | Round 2 | Round 3 | Convert | Round 4 | Round 5 | Round 6 | Round 7 | Self | Self | Round 8 | Total |
|---|---|---|---|---|---|---|---|---|---|---|---|---|
| Week of: | 8/24/20 | 9/7/20 | 9/21/20 | | 10/5/20 | 10/19/20 | 11/2/20 | 11/16/20 | | | 11/30/20 | |
| Positives | | | | | | | | | | | | |
| LS (1-6) | 1 | | 2 | 1 | 1 | | | 4 | 1 | | 4 | 14 |
| MS (7-9) | | | | | | | | 1 | | | 3 | 4 |
| US (10-12) | | 1 | | | | | 2 | | | | | 3 |
| Adults | | 1 | | | | | 1 | | 1 | 1 | | 4 |
| Total Positives | 1 | 2 | 2 | 1 | 1 | 0 | 3 | 5 | 2 | 1 | 7 | 25 |
| Total Tests | 743 | 1156 | 1366 | | 1370 | 1448 | 1417 | 879 | | | 1666 | |
